# COVID-19 and Curcumin: Using VOSviewer software to explore scientific landscapes – A bibliometric analysis

**DOI:** 10.1101/2022.04.10.22273674

**Authors:** Mostafa H. Abd El Wahab

## Abstract

**Background:** Curcumin is derived from Turmeric which is a spice with an old history in the oriental world. For this reason, it was a subject for continuous research over years and some studies reported preliminary positive results in arthritis and metabolic syndrome. When COVID-19 declared as a global pandemic, debates exploded again regarding its effects in mitigating deleterious effects of the viral infection. However, being a traditional remedy flooded readers with thousands of publications, some of these was scientifically rigid and others were fraudulent. We aim to use VOSviewer software to visualize scientific landscape in this topic, to highlight the trends and identify main supporting bodies.

**Methods:** We searched Web of Science (WOS) core collection database for publications between December 2019 and April 2022. Data collected include: year of publication, keywords, type of the document, author names, affiliations, abstracts and number of citations. VOSviewer 1.6.18 was used to analyze co-citation, co-occurrence, and publication trends. Analysis considered one weight attribute which is “total link strength attributes”.

**Results:** A total of 205 publications (N=205) were included in the analysis. Most studies were original research articles (50.7%). Mean citation count of the top 10 cited articles was 37.9 (range 22 and 111). Country of corresponding author of these 10 studies was India in 5 (50%), Iran in 3 (30%). Organizational analysis revealed 5 Iranian universities as being the main research bodies with total link strengths (TLS) of 100. Co-occurrence of keywords identified “viral inhibition, oxidative stress, molecular docking, NF-kB pathway” as the most frequent mentioned keywords. Trend analysis showed negative trend with less publications covering this topic, chronologically.

**Conclusion:** Curcumin resided within the oriental tradition for years, it is no surprise that main supporting bodies were oriental. VOSviewer provides an easy, user-friendly options to handle bibliographic data.

## 1. Introduction

The first cases of Severe Acute Respiratory Syndrome Coronavirus 2 (SARS-Cov-2) were reported in December 2019, soon after it was declared as global pandemic and international health crisis^1^. The pandemic placed heavy burdens on every healthcare system and many of these are already overwhelmed by huge number of patients^2^. In response to the growing challenges, literature regarding COVID-19 has expanded considerably.A study by Loannidis et al. based on data derived from Scopus database found that Till 1 August 2021, a total of 210,183 publication covering different aspects on COVID-19 were published and around 23520 authors of these publications were at the top 2% of their subfiled^3^. Submission to Elsevier’s magazines and periodicals alone increased by around 270.000 in 2020, which equals to 58% increase compared to the previous year, the increase was mainly related to expansion in medical field submissions^4^.

Turmeric is a spice with old history in the oriental world. Turmeric is derived from rhizomatous herbaceous perennial plant, which is a part of the ginger family^5^. Curcumin is derived mainly from Turmeric; it is also known as diferuloylmethane which is a naturally occurring polyphenol. Some in-vitro studies showed potential effect on modifying and modulating many signaling pathways^6^. However, its in-vivo abilities are still lacking much research, mostly due to its poor bioavailability resulting from protracted absorption kinetics, rapid metabolism and ultra-fast clearance^7^. Proposed benefits include improving systematic markers of inflammatory stress^8^, anti-arthritic effects^9^ and improving metabolic syndrome^10^.

However, being a traditional part of the oriental kitchen, it is often being mislabeled! Many myths are generated about its potentials for treating everything! The reader is often flooded with hundreds of publications and media outputs about its potential benefits. Analyzing the patterns governing these trends needs tremendous efforts, additionally, identifying its scientific truth and debunking myths is like grasping at straws. We think that bibliometric analysis can help solving these problems, by providing scooping review of the most cited articles, research bodies and publication dynamics, the reader is able to grasp the broadest lines of knowledge.

Bibliometric analysis is the statistical method which provides quantitative analysis of research papers concerned about one special topic and using mathematical methods^11^.In this study we aim to provide a bibliometric analysis of Curcumin publications in COVID-19 topic, we search Web of Science (WOS) – Core Collection database, this database provides almost all important research publications as well as strong analytical tools for bibliometric research. We use the VOSviewer software developed by van Eck and Waltman^12^. This software provides the capabilities of displaying large bibliometric maps in an easy-to-interpret way, with the additional advantages of user-friendly interface and open access with no charges.

### 2. Methods

We searched the global literature regarding Curcumin and COVID-19 virus within Web of Science (WOS) core collection database for publications between December 2019 and April 2022, Search took place on 10^th^ April 2022. This database requires a subscription, but for the Egyptian researchers it can be accessed freely through Egyptian Knowledge Bank (EKB) portal. Since search was done in public databases, no ethical approval is required. A single investigator extracted the data. Search terms for Curcumin were Curcumin OR Curcumine or “Turmeric yellow” OR “Curcuma Longa”. Details of search methodology are described in Appendix 1. Search was limited only for articles published in English. We used trend analysis function in WOS Core Collection database which provides insights about research bodies, countries and disciplines involved. Since no reporting guidelines for bibliometric analyses are included within EQUATOR NETWORK, we considered PRISMA statement 2020. Appendix 2 provides the items fulfilled in this manuscript.

The information collected include year of publication, keywords, type of the document, author, affiliation, abstracts and number of citations. Articles not fulfilling these requirements are typically excluded, all retrieved data were exported into a tab delimited file. VOSviewer 1.6.18 was used to analyze Co-citations, Co-occurrence and publication trends. Analysis considered one weight attribute which is “Total line strength attribute”.

## 3. Results

### 3.1 Bibliometrics of the output

A total of 206 publications were retrieved (N _initial_=206), quick title review excluded one study due to mismatch with inclusion criteria leaving a valid total of 205 publications (N _valid_ =205). 104 publications (50.7%) were original research articles, 91 (44.4%) were review articles and 10 (4.9%) were letters, book chapters or editorial materials. A total of 21 papers (10.2%) were published in 2022, 116 (56.6%) in 2021 and 68 publications (33.2%) in 2020. All manuscripts identified were written in English. A total of 165 publications (80.5%) were open access. Main research categories covered in Web of Science Core Database were pharmacology in 57 (27.8%), biochemistry in 30 (14.6%) and general chemistry in 26 (12.7%).

### 3.2 Bibliometric analysis of Citations

Top 10 cited studies are listed in Table (1), most of the studies were descriptive studies, review articles or clinical trials. Mean citations count was 37.9 with results ranging between 22 and 111. Country of corresponding author of the top 10 cited studies was India in 5 (50%), Iran in 3 (30%), Bangladesh and Egypt with one for each (10% each). For the total 205 studies, we performed an analysis considering authors, organizations, and countries of the studies separately. We identified top 5 in each analysis based on total link strengths (TLS). Organizational analysis of TLS revealed 5 Iranian universities having the highest TLS, with total TLS of 100. Country analysis, however, showed that Iran as a country lied second with TLS of 217, after India which came first with 248 TLS. Individual author analysis revealed Jamiaalahmadi, Tannaz as the most active author in the field of Curcumin and COVID-19 with TLS of 36, He was one of the coauthors who participated in the publication titled “Potential effects of curcumin in the treatment of COVID-19 infection” which is highest cited publication in our analysis. Table (2) summarizes these data, Figure (1) shows VOSviewer map based on Country, Figure 2 shows the data based on organization.

**Table (1).** Top 10 cited studies that examine relations between Curcumin and COVID-19

| **Citation Rank** | **Title** | **Country of corresponding author** | **Publication date** | **Total citations** |
| --- | --- | --- | --- | --- |
| 1 | Potential effects of curcumin in the treatment of COVID-19 infection | Iran | November 2020 | 111 |
| 2 | Nano-curcumin therapy, a promising method in modulating inflammatory cytokines in COVID-19 patients | Iran | January 2021 | 62 |
| 3 | Identification of phytochemicals as potential therapeutic agents that binds to Nsp15 protein target of coronavirus (SARS-CoV-2) that are capable of inhibiting virus replication | India | May 2021 | 32 |
| 4 | In silico Screening of Natural Compounds as Potential Inhibitors of SARS-CoV-2 Main Protease and Spike RBD: Targets for COVID-19 | India | Jan 2021 | 27 |
| 4 | Oral nano-curcumin formulation efficacy in management of mild to moderate hospitalized coronavirus disease-19 patients: An open label nonrandomized clinical trial | Iran | May 2021 | 27 |
| 6 | Bioactive Polyphenolic Compounds Showing Strong Antiviral Activities against Severe Acute Respiratory Syndrome Coronavirus 2 | Egypt | June 2021 | 26 |
| 7 | Virtual screening and molecular dynamics simulation study of plant-derived compounds to identify potential inhibitors of main protease from SARS-CoV-2 | Bangladesh | March 2021 | 24 |
| 7 | Calming the Storm: Natural Immunosuppressants as Adjuvants to Target the Cytokine Storm in COVID-19 | India | January 2021 | 24 |
| 7 | Antiviral and immunomodulatory activity of curcumin: A case for prophylactic therapy for COVID-19 | India | February 2021 | 24 |
| 10 | Oral Curcumin with Piperine as Adjuvant Therapy for the Treatment of COVID-19: A Randomized Clinical Trial | India | May 2021 | 22 |

**Table (2).** Ranks of organizations, countries and authors based on TLS

| **Ran**<br>**k** | **Organization** | **TLS/organizatio**<br>**n** | **Countr**<br>**y** | **TLS/Countr**<br>**y** | **Author** | **TLS/Autho**<br>**r** |
| --- | --- | --- | --- | --- | --- | --- |
| 1 | Mashhad University of Medical Sciences | 32 | India | 248 | Jamia Alahmadi, Tannaz | 36 |
| 2 | Tehran University of Medical Sciences | 30 | Iran | 217 | Sahebkar, Amirhossein | 36 |
| 3 | Tabriz University of Medical Sciences | 19 | USA | 182 | Ahmadi, Majid | 27 |
| 4 | Islamic Azad University | 13 | Brazil | 105 | Aslani, Saeed | 27 |
| 5 | Shahid Beheshti University | 6 | England | 102 | Roshanger, Leila | 27 |

**Figure 1.**
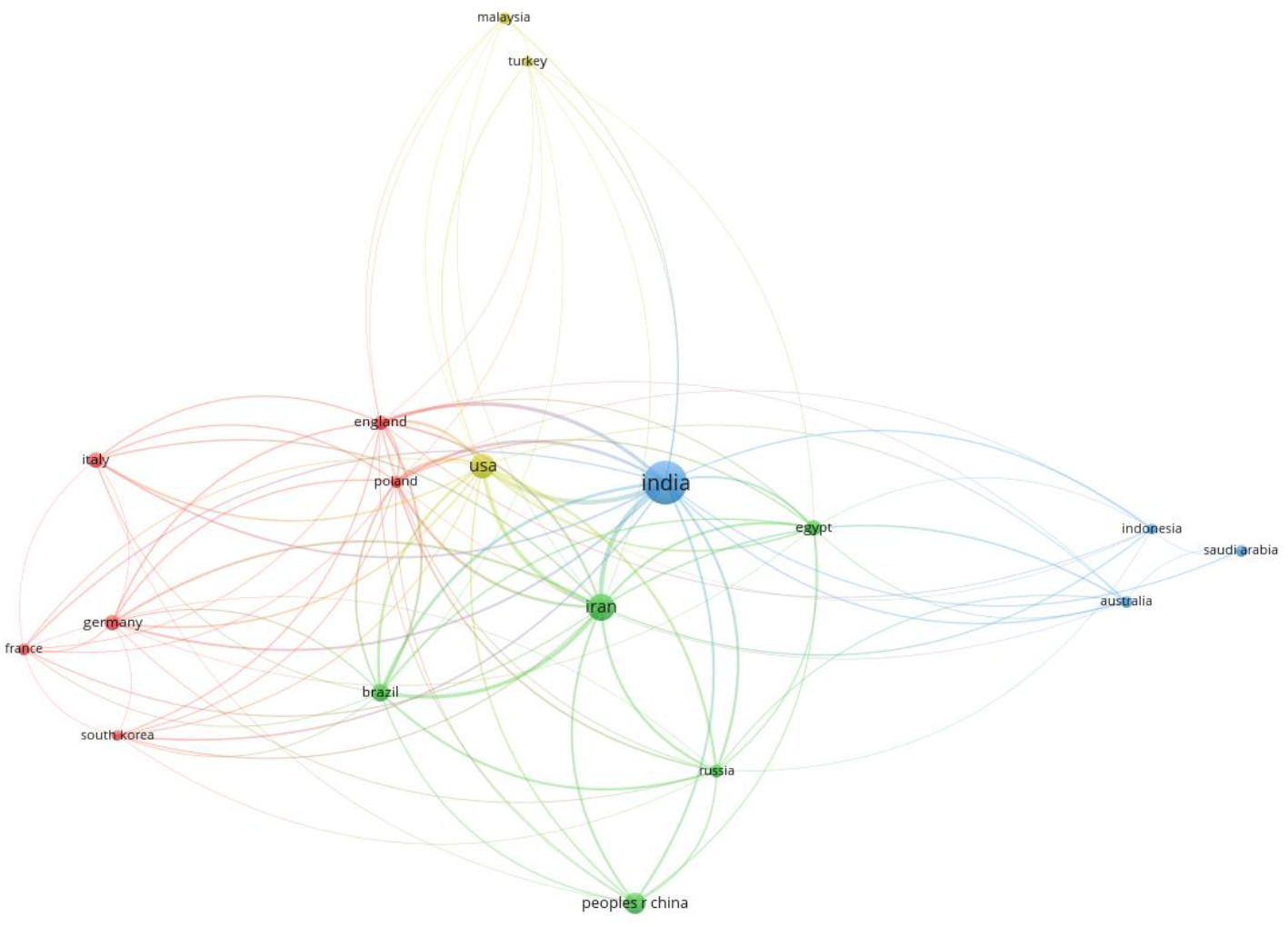
Authorship analysis based on the country, size of the nodes indicates the degree of contribution, curves represent links between different countries in authorship

**Figure 2.**
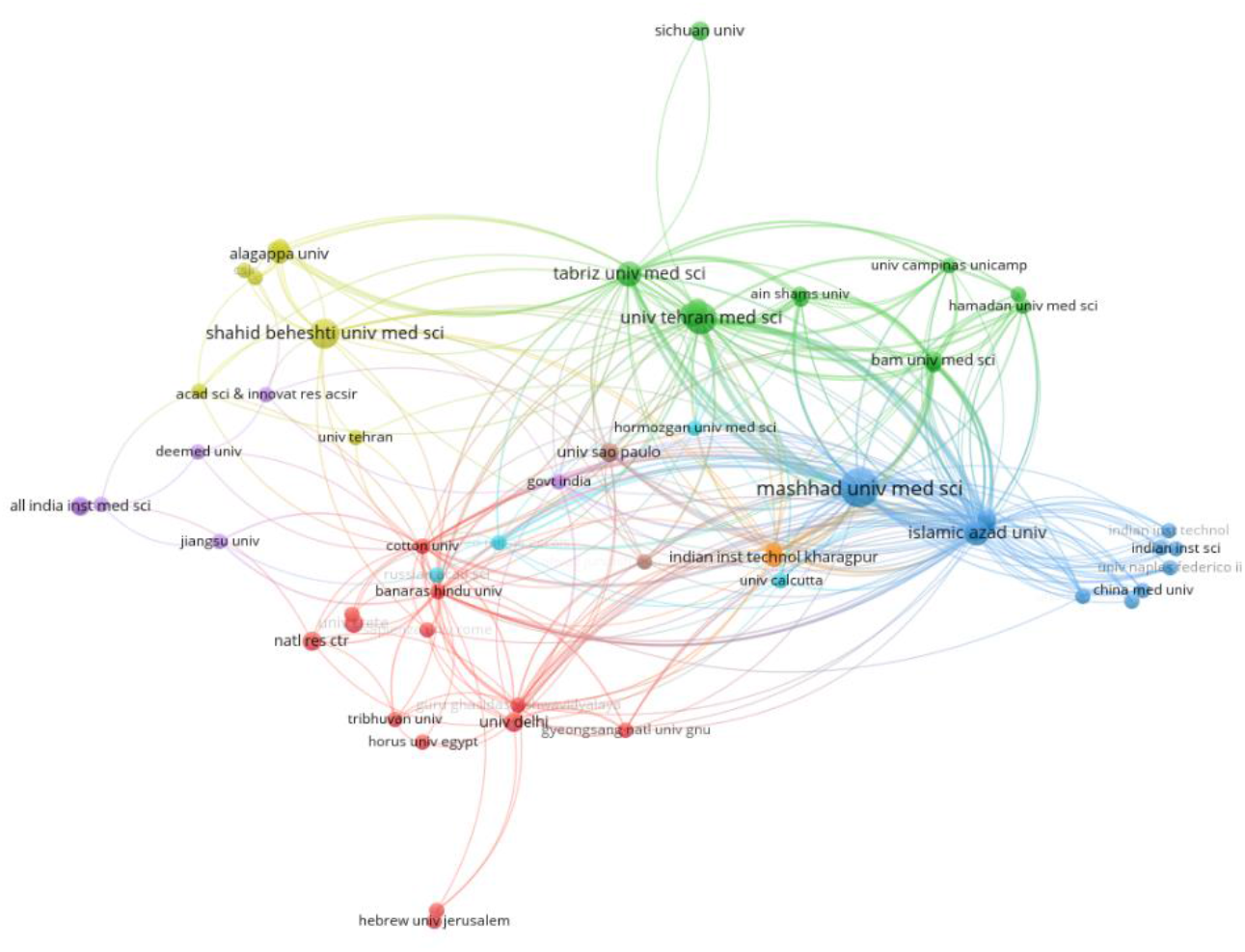
Authorship analysis based on organization, size of the nodes indicates the degree of contribution, curves represent links between different organizations

### 3.3 Bibliometric analysis of keywords

Regarding keyword analysis, VOSviewer software features two options, one for keywords provided by the authors and the second for keywords provided by authors in addition to others extracted from title and abstract. We choose the second option to catch changes as whole. Minimum number of occurrences of 5 in WOS database was considered in the analysis, of a total 1223 keywords, 65 met the threshold. We excluded formal keywords like (COVID-19, Curcumin). Important Keywords (in order of occurrence) were Inhibition of virus, oxidative stress, molecular docking, NF-kB pathway, antioxidant, viral expression, antiviral activity, nanoparticles, and Resveratrol.

### 3.4 Bibliographic analysis of the trends

Initially VOSviewer identified only 118 publications as being connected bibliographically togethers, this means that remaining 87 publications are not mentioned in any other studies within the same field. By default, VOSviewer asks you if you want dropping these items. After choosing this option the software tries to create clusters of publications that cover the same field, initial analysis revealed 16 different clusters with minimum of 5 in each cluster, this reflects large heterogeneity within the sub-domains of the research. We increased the minimum number of publications within each cluster to 15 to combine related sub-domains together, this identified 5 different areas of research. Figure (3) shows these clusters, the red cluster represents publications investigating curcumin as a potential treatment for modulating inflammatory cytokines, green cluster comprises publications investigating effects on Curcumin on virus binding and entry into cells. Blue cluster indicates publications that uses Curcumin as a direct treatment for COVID-19. Yellow cluster comprises publications that exploring characteristics of Curcumin that may need further studies and the purple cluster in concerned with its antioxidant effects.

**Figure 3.**
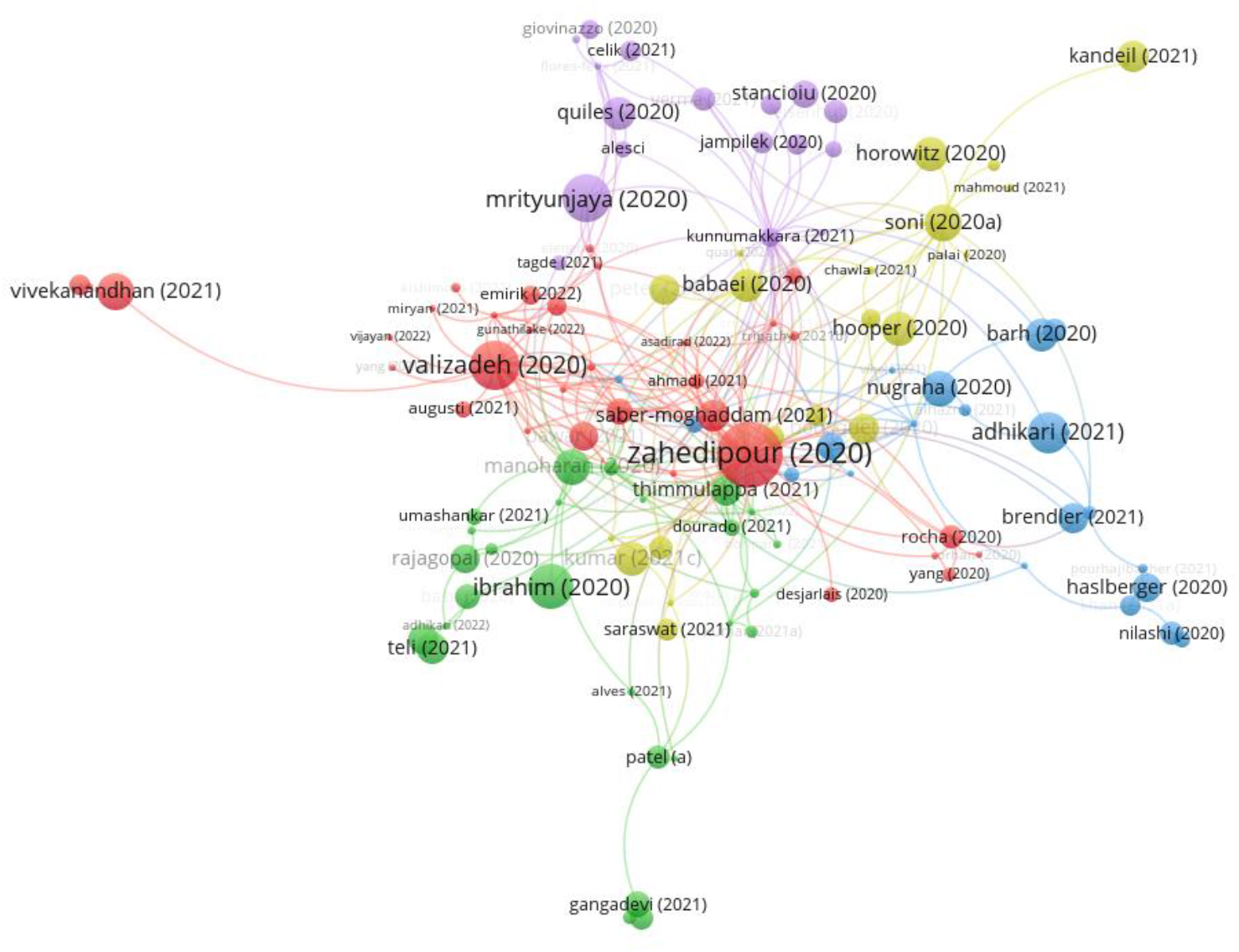
Clusters of different publications based on the domain it covers, size of the nodes indicates the degree of influence, curves represent citation relations between different publications

Overlay visualization option allows chronological view of publications, by default older studies are given purple color and recently published studies are given the yellow color. As seen in figure (4), the main bulk of research took place in the year 2020, however care should be taken in interpreting these results, as the weight of nodes are assigned based on citation volume, thus older studies are more likely to get cited than recently published ones. Despite this, the overall trend shows decline in the interest of Curcumin effects on COVID-19, many of the newest studies are meta-analyses of the trials examining the potential effects of Curcumin on reducing COVID-related mortality.

**Figure 4.**
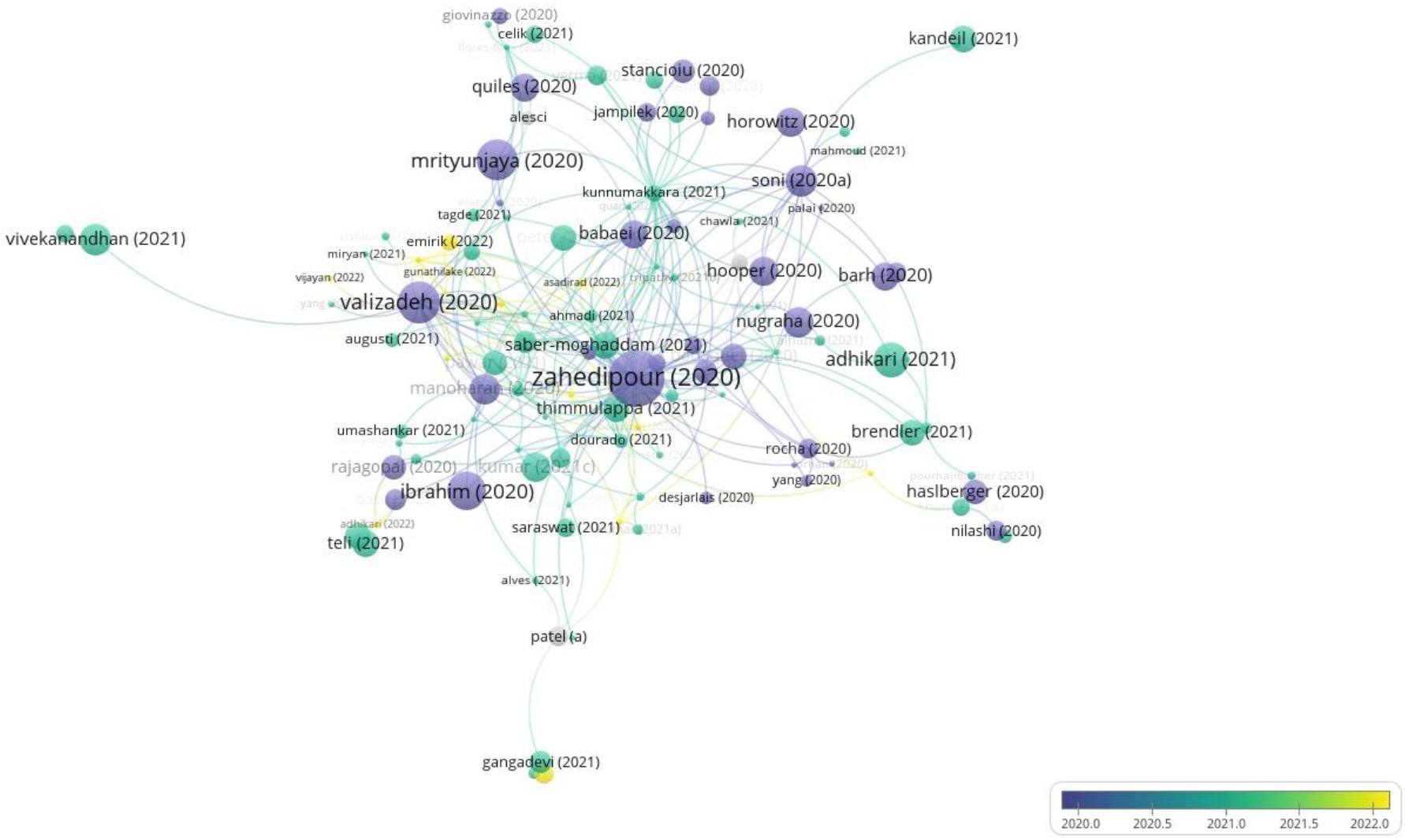
Bibliographic view of trends, older studies are colored in purple while new studies are colored yellow, as seen in the indicator. Size of the node represents its weight based on citation relations; lines represent citation relations between different nodes

## 4. Discussion

### 4.1 Why bibliometrics at all?

Bibliometric analyses provide a scooping overview of the research within a specific nosology, it is an instrument to guide decision making. Bormann and Daniel seek a deeper understanding of the motivations that led scientists to cite each other, they say: “citing behavior is not motivated solely by the wish to acknowledge intellectual and cognitive influences of colleague scientists, since the individual studies reveal also other, in part non-scientific, factors that play a part in the decision to cite^13^”. Robert Merton theory of citing behavior states that, by citation process, the authors acknowledge their peers, and thus they get rid of the conceptual burden of their guidance^14^. Bibliometrics, at least in part, shows the dynamics of thinking within a special context, it also reflects which ideas are prevailing within certain setting.

Bibliometric reviews also differ considerably from review articles. Review articles are built based on critically selected publications derived from certain scientific nosology. Professionals, who produced this content target their peers who know the field thoroughly, an external gazer will be left in the fog. Bibliometric analyses, on the other hand, are less specialized, from technical view of point. An external gazer will be able to catch main themes, new trends, competing groups and possibilities for cooperation^15^. This is particularly important in the current era, as medical practitioners are super-specialized with millions of publications out every year, still, medical practice requires a working knowledge of the surrounding subspecialities.

This review searched the effects on Curcumin on COVID-19. We identified a total of 205 publications. Top 5 organizations were Iranian universities with total link connections of 100. Iran as a country came second only after India, with total link connections of 217 and 248, respectively. This can be explained by the fact that curcumin is mainly an oriental spice, people deal with it every day, and they are obliged to think about it! However, this thinking wasn’t narrow. Research topics covered many fields, from its interaction with virus entry to the cells, to direct antiviral effects to its antioxidant functions. A plethora of ideas that bibliometric analysis made it possible for the reader!

### 4.2 Basic bibliographic terminologies

Bibliometric units are comprised of nodes and edges, nodes are any items that can be used as a unit of analysis (publications, journals, organizations, or countries). Edges represent the internal relation between these nodes. Many internal relations can be examined; however, most analyses focus on citation relations. If a certain publication cites two articles, those two articles are “Co-cited”. Bibliographic coupling is the opposite of co-citation, two publications can be bibliographically coupled when a third article is cited by both publications. Number of Co-occurrence of keywords refers to the number of publications into which those keywords were considered together^16^.

### 4.3 Your bibliographic analysis is your delicate search strategy

A good bibliographic analysis stems from a “winning literature search^17^”. First, a good research question should be considered. Choosing a broad research question will flood you with thousands of publications, choosing a narrow research question will minimize the catch. Choosing the suitable database is also of utmost importance, literature databases can be broadly divided into “white literature” which contains the peer-reviewed content and the “grey literature” which involves materials that may not be formally published by peer-reviewed journals. Databases vary considerable regarding the data available and types of file exporting capabilities.

### 4.4 VOSviewer, software overview

Many visualization techniques are available for visualizing bibliometric networks. VOSviewer uses a distance-based approach where the distance between two nodes indicates the degree of relatedness of those two nodes, this contrast with other visualization techniques as graph-based approaches, timeline-based approaches, and self-organizing maps. Publications usually have differences in magnitude compared to their counterparts, highly cited publications can have thousands of connections, and this makes visualization process difficult. VOSviewer targets these problems by a process called “normalization”. After normalization the software positions different nodes in a two-dimensional field that is projected as one of three options: network visualization, overlay visualization and density visualizations. Details of differences between these methods of display are beyond the scope of the review^18^.

## 5. Conclusion

As curcumin was popular in the oriental tradition for centuries, it is no surprise that most authors and research bodies that examined its effect on COVID-19 were oriental. VOSviewer software is very successful in viewing scientific landscape within certain nosology.

## Supporting information

Supplement 2, EQUATOR guidelines

Supplement 1, Search catch

## Data Availability

All data produced in the present study are available upon reasonable request to the authors

https://www.webofscience.com/wos/woscc/basic-search

## References

1 Wang C, Horby PW, Hayden FG, Gao GF. A novel coronavirus outbreak of global health concern. The Lancet. 2020 Feb 15;395(10223):470–3.

2 Emanuel EJ, Persad G, Upshur R, Thome B, Parker M, Glickman A, et al. Fair Allocation of Scarce Medical Resources in the Time of Covid-19. N Engl J Med. 2020 May 21;382(21):2049–55.

3 Ioannidis, J., Salholz-Hillel, M., Boyack, K. W., & Baas, J. (2021). The rapid, massive growth of COVID-19 authors in the scientific literature. Royal Society open science, 8(9), 210389. https://doi.org/10.1098/rsos.210389

4 Else, H. (2020). How a torrent of COVID science changed research publishing — in seven charts. Nature, 588(7839), 553–553. https://doi.org/10.1038/d41586-020-03564-y

5 Priyadarsini K. I. (2014). The chemistry of curcumin: from extraction to therapeutic agent. Molecules (Basel, Switzerland), 19(12), 20091–20112. https://doi.org/10.3390/molecules191220091

6 Gupta, S. C., Patchva, S., & Aggarwal, B. B. (2013). Therapeutic roles of curcumin: lessons learned from clinical trials. The AAPS journal, 15(1), 195–218. https://doi.org/10.1208/s12248-012-9432-8

7 Hewlings, S. J., & Kalman, D. S. (2017). Curcumin: A Review of Its Effects on Human Health. Foods (Basel, Switzerland), 6(10), 92. https://doi.org/10.3390/foods6100092

8 Sahebkar A., Serbanc M.C., Ursoniuc S., Banach M. Effect of curcuminoids on oxidative stress: A systematic review and meta-analysis of randomized controlled trials. J. Funct. Foods. 2015;18:898–909. doi: 10.1016/j.jff.2015.01.005

9 Henrotin, Y., Priem, F., & Mobasheri, A. (2013). Curcumin: a new paradigm and therapeutic opportunity for the treatment of osteoarthritis: curcumin for osteoarthritis management. SpringerPlus, 2(1), 56. https://doi.org/10.1186/2193-1801-2-56

10 Mohammadi, A., Sahebkar, A., Iranshahi, M., Amini, M., Khojasteh, R., Ghayour-Mobarhan, M., & Ferns, G. A. (2013). Effects of supplementation with curcuminoids on dyslipidemia in obese patients: a randomized crossover trial. Phytotherapy research : PTR, 27(3), 374–379. https://doi.org/10.1002/ptr.4715

11 Chen, C., Dubin, R., & Kim, M. C. (2014). Emerging trends and new developments in regenerative medicine: a scientometric update (2000 - 2014). Expert opinion on biological therapy, 14(9), 1295–1317. https://doi.org/10.1517/14712598.2014.920813

12 van Eck, N. J., & Waltman, L. (2010). Software survey: VOSviewer, a computer program for bibliometric mapping. Scientometrics, 84(2), 523–538. https://doi.org/10.1007/s11192-009-0146-3

13 Bornmann, L., & Daniel, H.-D. (2008). What do citation counts measure? A review of studies on citing behavior. Journal of Documentation, 64(1), 45–80.

14 Merton, R. K. (1973). Priorities in scientific discovery. In R. K. Merton (Ed.), The sociology of science: Theoretical and empirical investigations (pp. 286–324). Chicago: University of Chicago Press.

15 Ellegaard, O., & Wallin, J. A. (2015). The bibliometric analysis of scholarly production: How great is the impact?. Scientometrics, 105(3), 1809–1831. https://doi.org/10.1007/s11192-015-1645-z

16 Eyers J. E. (1998). Searching bibliographic databases effectively. Health policy and planning, 13(3), 339–342. https://doi.org/10.1093/heapol/13.3.339

17 Ecker, E. D., & Skelly, A. C. (2010). Conducting a winning literature search. Evidence-based spine-care journal, 1(1), 9–14. https://doi.org/10.1055/s-0028-1100887

18 Van Eck, N.J., & Waltman, L. (2014). Visualizing bibliometric networks. In Y. Ding, R. Rousseau, & D. Wolfram (Eds.), Measuring scholarly impact: Methods and practice (pp. 285–320). Springer

