## Supplement 1, Search catch for "COVID-19 and Curcumin: Using VOSviewer software to explore scientific landscapes – A bibliometric analysis"

| Search Terms | #1 “Corona virus” OR COVID-19 OR “SARS-Cov-2” OR “severe acute respiratory syndrome coronavirus2” OR nCoV-2019 OR “Wuhan AND Corona virus”  #2 Curcumin OR Curcumine OR “Turmeric yellow” OR “Curcuma longa”  Final search: #1 AND #2 |
| --- | --- |
| Search history | 10-4-2022 |
| Database | Web of Science – Core Collection |
| Total results | 206 |
| Total publications retrieved | 205 |
| Name of researcher | Mostafa H. Abd El Wahab |
